# Accuracy of CSF Tap Test and Lumbar Infusion Test in Predicting Shunt Response in Idiopathic Normal Pressure Hydrocephalus: A Systematic Review and Meta-Analysis

**DOI:** 10.1101/2025.11.09.25339846

**Authors:** Aminu Aliyar, Deepa Dash, Alfonso Fasano, Manya Prasad, Aparna Wagle Shukla, Ashish Dutt Upadhyay, Soaham Desai, Sagar Poudel, Pramod Kumar Pal, Arunmozhimaran Elavarasi

## Abstract

**Background:** Idiopathic normal pressure hydrocephalus (iNPH) presents with gait disturbance, cognitive impairment, and urinary incontinence. The cerebrospinal fluid tap test (CSF-TT) and lumbar infusion test (LIT) are commonly used to predict postoperative improvement after shunt surgery; however, their validity remains debated.

**Methods:** We performed a systematic review and meta-analysis to assess the sensitivity and specificity of CSF-TT and LIT. The protocol was registered in PROSPERO (CRD42023454502). Reporting followed the PRISMA guidelines, and certainty of evidence was assessed using the GRADE approach.

**Results:** From 1762 studies, 14 were included, reporting 697 CSF-TT and 393 LIT patients with shunt surgery as the reference standard. Considerable heterogeneity existed in test protocols, timing of assessment, and outcome measures. Several studies were retrospective with a high risk of bias. Using a bivariate random-effects model, pooled sensitivity and specificity of CSF-TT were 67.5% (95% CI 52.2–79.8, I^2^ 82.3%) and 53.3% (40.7-65.4, I^2^ 49.4%), respectively. The certainty of evidence was very low for sensitivity due to bias, inconsistency, and imprecision, and low for poor specificity due to bias and inconsistency. LIT showed a pooled sensitivity of 81% (70.3–88.3, I^2^ = 27.6%) and specificity of 42.8% (20.8–68.1, I^2^ = 60.8%), with moderate certainty for sensitivity and poor specificity.

**Conclusion:** Both CSF-TT and LIT demonstrate only modest accuracy in predicting shunt outcomes. The pooled specificity of the CSF-TT is similar to a coin toss, limiting its standalone clinical utility. The moderate certainty regarding the poor specificity of the LIT highlights the need for improved prognostic models. These tests should be interpreted in conjunction with other clinical and imaging findings, rather than in isolation. We need standardized, high-quality studies to better define their diagnostic value and support shared decision-making.

## Background

Normal pressure hydrocephalus (NPH) was first described by Hakim and Adams in 1965. It is characterized by the triad of gait disorder, cognitive impairment, and urinary incontinence. The prevalence of possible idiopathic NPH (iNPH) ranges from 10/100,000 to 29/100,000, with age-specific rates varying from 3.3/100,000 in individuals aged 50-59 to 5.9% in those aged 80 years or older.^1^ In the right clinical setting, neuroimaging is done to demonstrate non-obstructive hydrocephalus. Several imaging features, such as the callosal angle, Evan’s index, and disproportionately enlarged subarachnoid spaces (DESH), have been described, which help identify patients for ancillary testing. Suspected patients undergo a CSF tap test (CSF-TT) and/or lumbar infusion test (LIT), with serial monitoring of gait scores and cognitive function. Improvement in symptoms is considered a harbinger of successful symptom control following CSF shunting, such as ventriculoperitoneal (VP) or lumbo-peritoneal shunting. External lumbar drainage (ELD) is another minimally invasive method for CSF diversion; however, since it requires hospital admission and carries some risks (e.g., infection), it is not commonly performed in the preoperative phase.

Guidelines suggest that these tests have higher specificity and lower sensitivity in predicting outcomes, with conflicting opinions regarding their predictive power and validity.^2^ While some guidelines acknowledge that the CSF-TT has poor sensitivity,^3,4^ they claim that its specificity is quite high, suggesting that in those with a positive tap test, the likelihood of clinical improvement following the surgery is high.^4^ The Japanese guidelines advise clinicians that even if ancillary tests are negative, patients may still be referred for shunt surgery if other criteria, such as clinical and radiological features, are consistent with the diagnosis. Despite these recommendations, there is a wide variation in clinical practice, and several institutions, such as the UK-NHS^5^ and the Canadian Health System^6^ use the CSF-TT results to decide who undergoes a definitive surgical procedure. The optimal cutoffs and timing of assessment following the CSF-TT to predict improvement after shunting are not known.^7,8^ Despite variable endorsement in international guidelines, evidence supporting standardized protocols for these tests remains limited. Since there is clinical equipoise regarding the utility of these tests, this systematic review aims to synthesize the diagnostic accuracy of CSF-TT and LIT for predicting shunt response in iNPH, with a focus on methodological heterogeneity and risk of bias across studies

## Methods

We conducted a systematic review and meta-analysis to investigate the sensitivity and specificity of CSF-TT and LIT in predicting outcomes following shunt surgery in patients with iNPH. The protocol for this review was registered in PROSPERO, with the registration number CRD42023454502. We adhered to the PRISMA guidelines in reporting this systematic review.

We included studies of patients with probable or possible NPH, defined according to clearly described clinico-radiologic criteria in the individual studies, who underwent diagnostic testing with a large-volume CSF-TT or LIT, followed by VP or lumboperitoneal shunt surgery, regardless of the results of these preoperative ancillary tests. These studies had reported clinico-radiological details and/or functional outcome of patients after surgery and were published in the English language. To avoid circular reasoning, we excluded studies that reported results of surgery in patients who were operated on based on the results of predictive tests (i.e., CSF-TT or LIT) and those that had incomplete clinico-radiological details or lacked objectively quantified outcomes following surgery. We developed a search strategy that included keywords and various terms to cover all aspects of iNPH. The search strategy was approved by all authors, following several iterations to finalize the approach (Appendix 1). We searched MEDLINE (Ovid), Embase, Cochrane CENTRAL, and conducted a PubMed search for studies not yet indexed in MEDLINE on October 22, 2025. We also searched the reference lists of all included studies and relevant systematic reviews for additional references, as well as Scopus databases for studies that fulfilled the inclusion criteria listed above. All studies obtained through electronic search were stored in Zotero reference management software (version 6.0.36, Corporation for Digital Scholarship, Virginia, USA) and then imported into Covidence systematic review software (Veritas Health Innovation, Melbourne, Australia), where the references were de-duplicated, screened, full text review, data extraction, and quality assessment were done. A pair of reviewers (AA and AE) independently screened titles and abstracts, reviewed the full texts of potentially eligible studies to determine the final eligible studies, and abstracted data. We abstracted the last name of the first author, year of publication, country, and hospital, as well as population, interventions, and outcomes, into prespecified abstraction forms. Disagreements between the reviewers were resolved by discussion or by a third reviewer (SD).

### Risk of bias assessment

QUADAS-2 (Quality Assessment of Diagnostic Accuracy Studies 2) framework^9^ was employed to assess the risk of bias. It was explored across four domains: patient selection (3 questions), index test (2 questions), reference standard (2 questions), and flow and timing (4 questions), which were answered as “yes,” “no,” or “unclear” based on the data reported in the individual studies. The pair of reviewers (AA and AE) assessed the risk of bias based on the answers to the questions as follows: low risk if all answers were ‘yes’; unclear risk of bias if at least one question was answered as ‘unclear’; and high risk of bias if at least one question was answered as ‘no.’ For the first three domains, the applicability concerns were assessed as low risk, unknown risk, or high risk if the population, index test, or reference standard did not align with the review question. Disagreements between the reviewers were resolved through discussion and, in some cases, by the third reviewer (SD).

### Thresholds for index test positivity and Definitions for the reference standard

The thresholds and definitions used in the individual studies were used for the purpose of analyzing the diagnostic accuracy.

### Statistical analysis

Sensitivity and specificity analyses were performed by extracting the number of patients with and without a response to the CSF-TT or LIT, as well as the number of patients who improved and those who did not improve following surgery. We conducted a meta-analysis of diagnostic test accuracy using the metadta command in Stata version 17.0 (StataCorp LLC, College Station, TX, USA). This command implements a bivariate random-effects model, which jointly analyzes sensitivity and specificity while accounting for their correlation and between-study variability.^10^ True positives (TP), false positives (FP), false negatives (FN), and true negatives (TN) were extracted from each study to estimate pooled sensitivity, specificity, diagnostic odds ratio, and construct the summary receiver operating characteristic (SROC) curve. The τ^2^ statistic and I^2^ were used to assess heterogeneity. No prespecified sensitivity analysis was planned regarding risk of bias assessment. A post hoc sensitivity analysis was performed by excluding studies that used different cutoffs for test positivity.

### Certainty of Evidence

We used the GRADE (Grading of Recommendations Assessment, Development and Evaluation) approach to assess the certainty of evidence for each primary outcome, considering factors such as risk of bias, inconsistency, indirectness, imprecision, and publication bias. To address grading inconsistency and imprecision, we employed a diagnostic threshold of 70% for both sensitivity and specificity, considering the clinical usefulness of the test.

## Results

The electronic search identified 1762 studies, and 452 duplicates were removed. One thousand three hundred and ten articles were screened for eligibility as per our inclusion and exclusion criteria, and 84 studies were selected for full-text review. We extracted data from 14 studies that reported the results of the index tests and reference standards as per the review question (Figure 1). Nine studies reported the diagnostic accuracy of the CSF-TT, and six studies reported the diagnostic accuracy of LITs in NPH.

**Figure 1.**
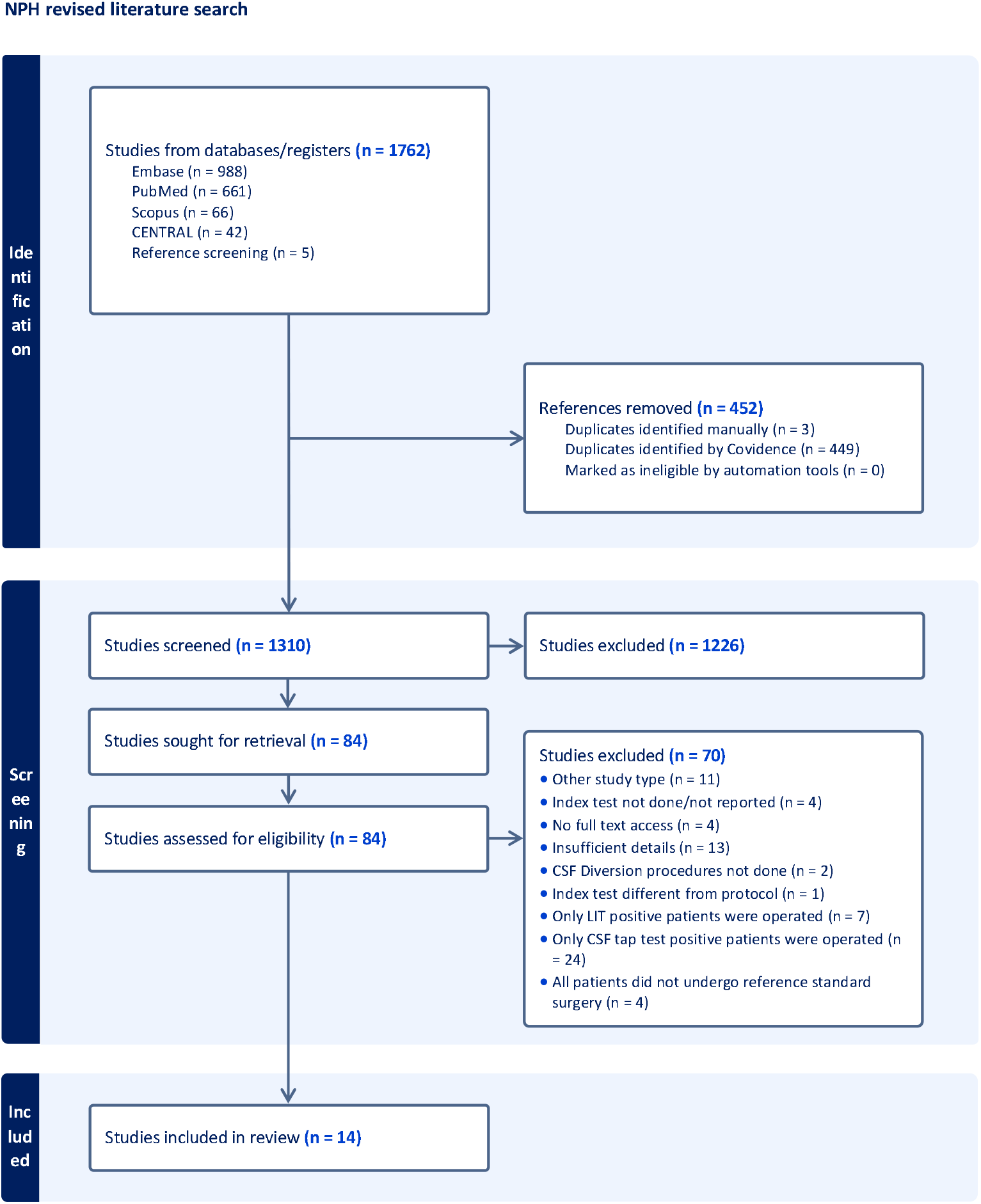
PRISMA flow diagram of study selection process

### CSF tap test and post-shunt outcomes

We identified nine cohort studies,^11–19^, comprising three prospective, four retrospective, and two with an unknown direction of data collection. A total of 697 patients were included. The characteristics of these studies have been described in detail in Supplementary Table 1. The mean ages of the patients ranged from 67 to 78.8 years. Gait dysfunction varied from 84 and 100%, urinary dysfunction between 67 and 82% and cognitive dysfunction between 72 and 84%. The duration of symptoms ranged from one month to 11 years in five studies; however, in four studies, the duration of symptoms was not reported. The radiologic findings, selection criteria for shunting, and the duration from evaluation to the therapeutic procedure varied significantly among the different studies, as shown in Table 1.

**Table 1:** CSF tap test: Technique, Assessment, tap responsiveness, shunt responsiveness, and diagnostic accuracy.

| Study ID; Country | CSF tap test procedure and criteria for positivity | Timing of surgery, outcome measures, and criteria for shunt responsiveness | Diagnostic accuracy measures |
| --- | --- | --- | --- |
| 1. **Wikkelso 1986; Sweden**<sup>11</sup> | CSF tap test procedure: lumbar puncture of 50 cc CSF<br>Tests: Psychometric testing, number of steps to walk 18 m<br>Assessment: Before the tap test and 2 hours after the tap test<br>Criteria for a positive tap test: The assessment was conducted 2 hours after the tap test. Identical forms 25 %; Bingley's memory test 25%; reaction time test 5%; and walking test 5% | Timing of shunt surgery after CSF tap test: NR<br>The postoperative assessment was done 3-6 months after shunt placement.<br>Criteria for shunt response: NR | Walk test: Sensitivity: 89; Specificity: 100<br>Memory test: Sensitivity: 77; Specificity: 75 |
| 2. **Ishikawa, 2012; Japan**<sup>12</sup> | CSF tap test procedure: A lumbar tap with the removal of 30 ml of CSF<br>Tests: iNPH grading scale; the Mini-Mental State Examination (MMSE), and the 3-meter timed 'Up & Go' test (TUG).<br>Assessment: Change in gait 1-2 days after tap, cognition, and urination at 1 week after tap<br>Criteria for a positive tap test: An improvement of one point or more on the iNPH grading scale (in each domain and its total), a more than 10% improvement in time on the TUG, or a more than 3-point improvement in the MMSE; Improvement in any of the total scores of iNPHGS, TUG, or MMSE was defined as positive with an additional variable of Tap-any | Timing of shunt surgery after CSF tap test: within 2 months<br>Assessment: at 12 months<br>Shunt responder: Improvement of one point or more on the modified Rankin scale | Gait scale: Sensitivity: 51.3<br>Specificity: 80; TUG<br>Sensitivity: 34.2 Specificity: 73.6<br>Urinary scale<br>Sensitivity 37.5 Specificity: 85<br>MMSE<br>Sensitivity 63.8 Specificity 30<br>Total gait scale Sensitivity 71.3 Specificity 65. Tap any<br>Sensitivity 92.5 Specificity 20 |
| 3. **Wikkelso, 2013**<sup>13</sup> | CSF tap test procedure: A CSF TT of 50 ml was performed at approximately 09:00h.<br>Tests: Gait - number of steps and the time needed to walk 10 m at free speed.<br>Psychometric performance using the Grooved Pegboard (Lafayette Instrument Co.), which measures manual dexterity as the time to fit 25 pegs into holes with randomly positioned slots, and the Stroop Test.<br>Assessment: Three hours after drainage of cerebrospinal fluid<br>Criteria for a positive tap test: Response to the CSF TT was expressed as the mean percentage change in all motor and psychometric tests compared with the results achieved 24 hours previously (the day before the combined CSF dynamic test). A mean increase of 5% was considered significant. | Timing of shunt surgery after CSF tap test: NR<br>Assessment: at 12 months postoperatively<br>Shunt responder: Improvement in the iNPH Scale was defined as an increase of 5 points or more, and in the mRS as a decrease of 1 point. | Improvement in motor and psychometric scores(>5%)<br>Sensitivity: 52%<br>Specificity: 59% |
| 4. **Yamada, 2017; Japan**<sup>14</sup> | CSF tap test procedure: 30 mL of CSF via a lumbar puncture.<br>Tests: iNPH grading scale, timed 3-m up-and-go test (TUG), and 3-m reciprocating walking test (RWT)<br>Assessment: Gait was evaluated within 24 hours, and cognitive impairment within 7 days<br>Criteria for positive tap test: Clinical improvement in gait was assessed by $\geq 1$ point improvement of the gait domain of the iNPHGS, and combined gait evaluation was defined as $\geq 1$ point improvement of the gait domain of the iNPHGS combined with $\geq 10\%$ shortening of the time on the TUG or 3-m RWT<br>Clinical improvement of cognitive impairment was assessed as $\geq 1$-point improvement in the cognitive domain of the iNPHGS. A combined cognitive evaluation was defined as a $\geq 1$-point improvement in the cognitive domain of the iNPHGS, combined with a $\geq 3$-point improvement on the MMSE or a $\geq 2$-point improvement on the FAB.<br>In addition, $\geq 1$-point improvement in the modified Rankin Scale and the total iNPHGS was used to assess general improvement. | Timing of shunt surgery after CSF tap test: within 3 months<br>Assessment: 12 months<br>Criteria for shunt responsiveness: improvement of the outcome variables at 12 months after the surgery. | iNPHGS Sensitivity: 61.3%<br>19/31; Specificity: 64.7%<br>33/51<br>mRS<br>Sensitivity 27.5<br>Specificity 93.3 |
| 5. **Liu 2021; China**<sup>15</sup> | CSF tap test procedure: 30 ml of CSF was removed via lumbar tap.<br>Assessment: Before and 8, 24, 72 h after LP<br>Test: (1) walking tests: 10-meter walking time, 10-meter walking steps, and 5-meter timed up and go test (TUG) time.<br>For the TUG, the amount of time required for a patient to rise from an armchair, walk 5 m, turn, walk back to the chair, and sit down again, at maximum speed was recorded. For 10Ti and 10St tests, the patient was also asked to walk at maximum speed. Each assessment was performed twice, and the average time and steps were calculated.<br>Cognitive assessment: the brief executive battery included the Symbol-Digit Modalities test, Trail Making Test A, and STROOP-C. The composite z-scores of the brief executive function battery were calculated by averaging the z-scores of the three tests according to the Chinese norm.<br>Criteria for positive tap test: CSF TT responders: (1) the time or number of steps in the walking tests were reduced by 10% at a minimum of one-time point after the CSF TT; or (2) the executive function composite z score was improved by more than 20% (Any time after tap test 8 h, 24 h and 72 hours) | The shunt was done within one month of TT.<br>Assessment: 34±26 months<br>Criteria for shunt responsiveness: Improvement reported by the patient or caregiver, accompanied by at least a one-point increase or more in the Japanese iNPHGS. | 24 h testing: TUG Sensitivity 35.7%; Specificity 66.7%,<br>10 m time test Sensitivity 37.5%, specificity 85.7%,<br>Composite Z score: Sensitivity 72.7%, Specificity: 80%,<br>Cumulative TUG sensitivity 57%, Specificity 43%,<br>Composite Z score sensitivity 82%, Specificity 60% |
| 6. Rydja 2021; Sweden<sup>16</sup> | CSF tap test procedure: A neurologist performed the LP with the patient in a lateral position. A spinal fluid manometer was used to measure the lumbar cerebrospinal fluid (CSF) pressure for 30 seconds before the removal of CSF.<br>Assessment: Approximately 3 hours after the LP<br>Tests: Gait and balance domains in the iNPH scale.<br>Criteria for positive tap test: Outcomes assessed 3 hours after the tap test. CSF TT responders with a positive outcome ($\geq 5$ points) in the gait domain on the Hellstrom iNPH scale | Surgery was done 126 (102-159) days after baseline assessment.<br>Assessment: 3 months following surgery.<br>Criteria for shunt responsiveness: A limit of $\geq 5$ points for the total Hellstrom iNPH scale is proposed as an improvement after surgery | Gait domain iNPH scale as the outcome in TT and total iNPH as postoperative outcome - Sensitivity: 68.1%; Specificity: 52%<br>Gait domain as an outcome in the CSF TT and the same outcome postoperatively: Sensitivity 72%, Specificity 52% |
| 7. Kameda, 2022; Japan<sup>17</sup> | CSF tap test procedure: 30 mL of CSF was tapped.<br>Assessment: If the tap test was performed as an inpatient investigation, the TUG assessment was mainly performed 1 or 3 days after the tap test. If the tap test was performed as an outpatient investigation, the TUG was assessed immediately after the tap test and at the next outpatient visit, up to 2 weeks later. If TUG was evaluated multiple times after the tap test, the best one was used.<br>Tests: The Functional Gait Assessment (FGA) is a 10-item evaluation of gait. Following the tap test, patients were asked to indicate whether they thought there was a change in their gait using a Global Rating of Change (GRC) scale.<br>Criteria for positive tap test to calculate diagnostic accuracy measures: FGA improvement of 4 or more, GRC improvement of 2 or more, TUG improvement of 10% or more | The timing of the LP shunt after the Tap test is not mentioned<br>Criteria for shunt responsiveness: Improvement in the Japanese iNPH score, cutoffs, and timing of assessment post-surgery are not mentioned. | TUG: Sensitivity 23%, specificity 71%<br>FGA Sensitivity 83% Specificity 17%<br>GRC 70%; Specificity: 25%<br>Missing outcomes in FGA and GRC |
| 8. vanBilsen 2022; Netherlands<sup>18</sup> | CSF tap test procedure: 50 mL tap test;<br>Criteria for positive tap test: No cutoffs for tap test responsiveness have been reported | The timing of surgery after the index test was not mentioned. Median follow-up duration was 6 months.<br>Improvement after shunting was defined as a minimum one-point improvement in the NPH score.<br>Median time to surgery: 1 month in responders (1-2); 1.5 months in non-responders (1-3.5) | Sensitivity: 77<br>Specificity: 25 |
| 9. Gao 2025; China<sup>19</sup> | CSF tap test procedure: Not reported<br>Assessment: gait was assessed using the 10 m walk test; Cognition was assessed with the mini-mental state examination; Urinary incontinence was subjectively assessed by patients on a scale from 0 to 10, indicating the severity from normal to severe. An identical assessment was performed 24–72 h later following the tap test.<br>Criteria for positive tap test: At least one of the following: (a) $\geq 20\%$ improvement in time or steps of the 10 m timed walk test and/or (b) $\geq 10\%$ improvement in MMSE score and/or (c) an improvement of $\geq 1$ point in Urinary Incontinence score. | Timing of surgery after the index test: Not reported<br>Median follow-up duration: 6-12 months<br>Criteria for shunt responsiveness: Krauss index at a follow-up of 6–12 months, where responders were defined as those with a Krauss index $\geq 0.5$. | Sensitivity: 63.5%<br>Specificity: 60% |

The clinical protocols applied for the tap test varied widely, and the clinical tests used were different and have been described in detail in Table 1. The volume of CSF tapped during the tap test ranged from 30 mL (Ishikawa et al.^12^, Yamada et al.^14^, Liu et al.^15^ and Kameda et al.^17^) to 50 mL (Wikkelsö et al.^11^, Wikkelsö et al.^13^, and van Bilsen et al.^18^). Improvements were evaluated as early as 2 hours after the tap (Wikkelsö et al.)^11^ to as late as 1 week for cognitive and urinary symptoms (Ishikawa et al.^12^; Yamada et al.^14^). Liu et al.^15^ performed serial assessments at 8, 24, and 72 hours.

Several different clinical assessment tools 10-meter walking test, Bingley’s memory test, reaction time test, MMSE, timed 3-m timed up-and-go test (TUG) and 3-m reciprocating walking test (RWT), iNPH grading system (iNPHGS), Grooved Pegboard (Lafayette Instrument Co Lafayette, IN), Stroop Test, Symbol-Digit Modalities test, and Trail Making Test A were used across various studies. Van Bilsen et al.^18^ did not report the criteria for a positive tap test, though they reported the number of patients with a positive tap test. The timing of shunt surgery after tap tests was either not mentioned or variable between the included studies. Similar heterogeneities existed in the criteria used to define improvement and the timing of assessing post-surgical outcomes (Table 1). The CSF-TT was not employed in the decision-making process in any of these studies.

We found the pooled sensitivity of the CSF-TT to be 67.5% (95% CI 52.2-79.8) with a substantial between-study variance and heterogeneity (τ^2^ 0.82, I^2^ 82.2%) and the pooled specificity to be 53.3% (40.7-65.5) with a moderate between-study variance and heterogeneity (τ^2^ 0.39, I^2^ 49.4%), (Figure 2) as also depicted by the (sROC) summary receiver operator characteristic curve (Supplementary Figure 1). The correlation between the logit-transformed sensitivity and specificity was -1, implying a perfect negative correlation, which is quite extreme and may indicate a strong threshold effect. This suggests that studies with higher sensitivity tend to have lower specificity, and vice versa.

**Figure 2.**
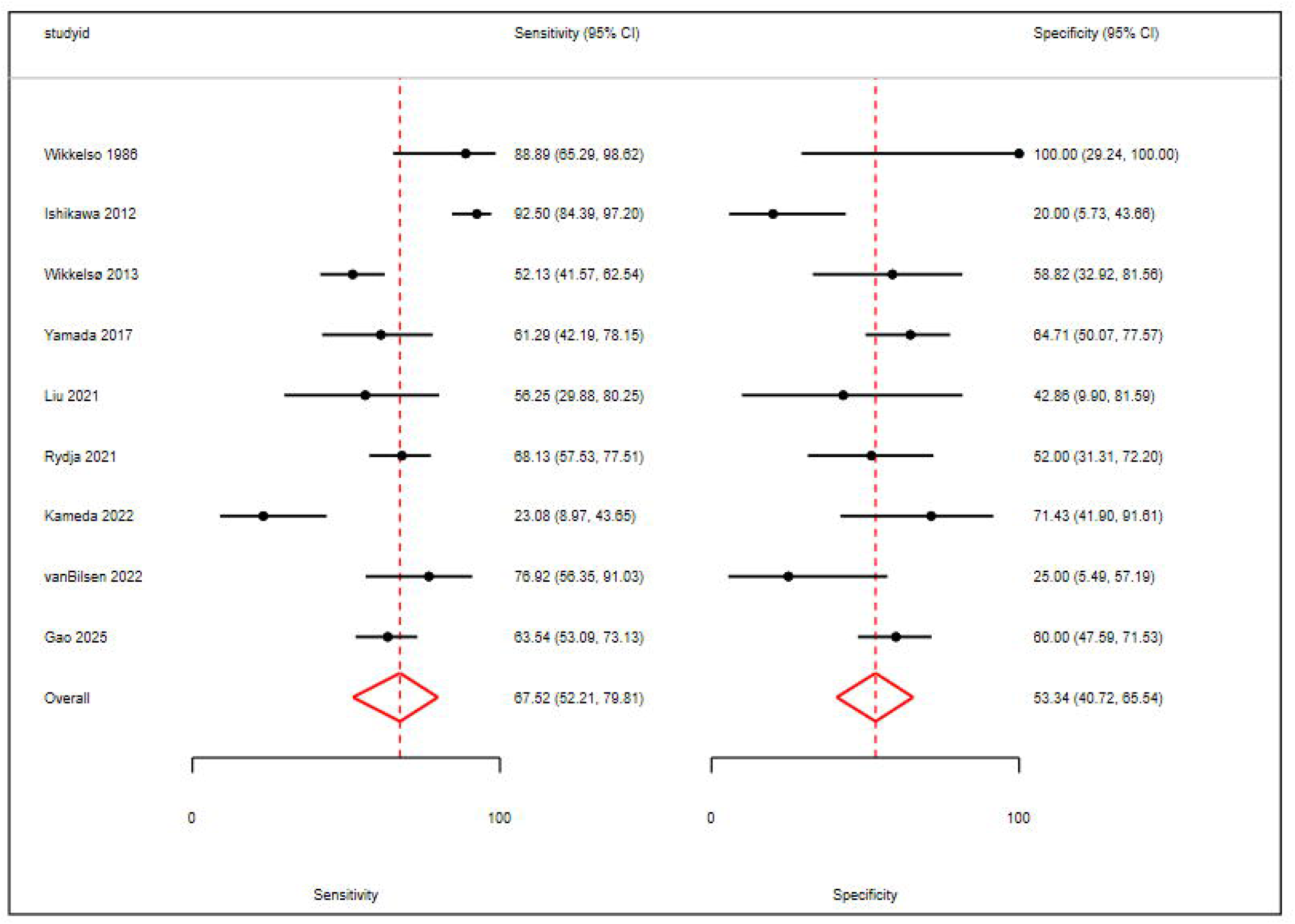
Forest plot showing pooled sensitivity and specificity of the CSF tap test (CSF-TT)

### Lumbar infusion tests and post-shunt outcomes

Six studies with 393 patients were included.^13,20–24^ Three studies were prospective cohort studies, two were retrospective cohort studies, and one had an unknown direction of data collection. The details of these studies are described in Table 2 and Supplementary Table 2. The mean ages ranged from 55 to 87 years. Only one study (Sorteberg et al).^22^ reported the prevalence of clinical symptoms, with gait dysfunction observed in 94% and urinary dysfunction in 88.2% of patients. The remaining studies did not provide data on the proportion of individuals with gait, urinary, or cognitive impairment. Similar to the studies reporting CSF-TT, the protocols varied widely among studies.

**Table 2:**
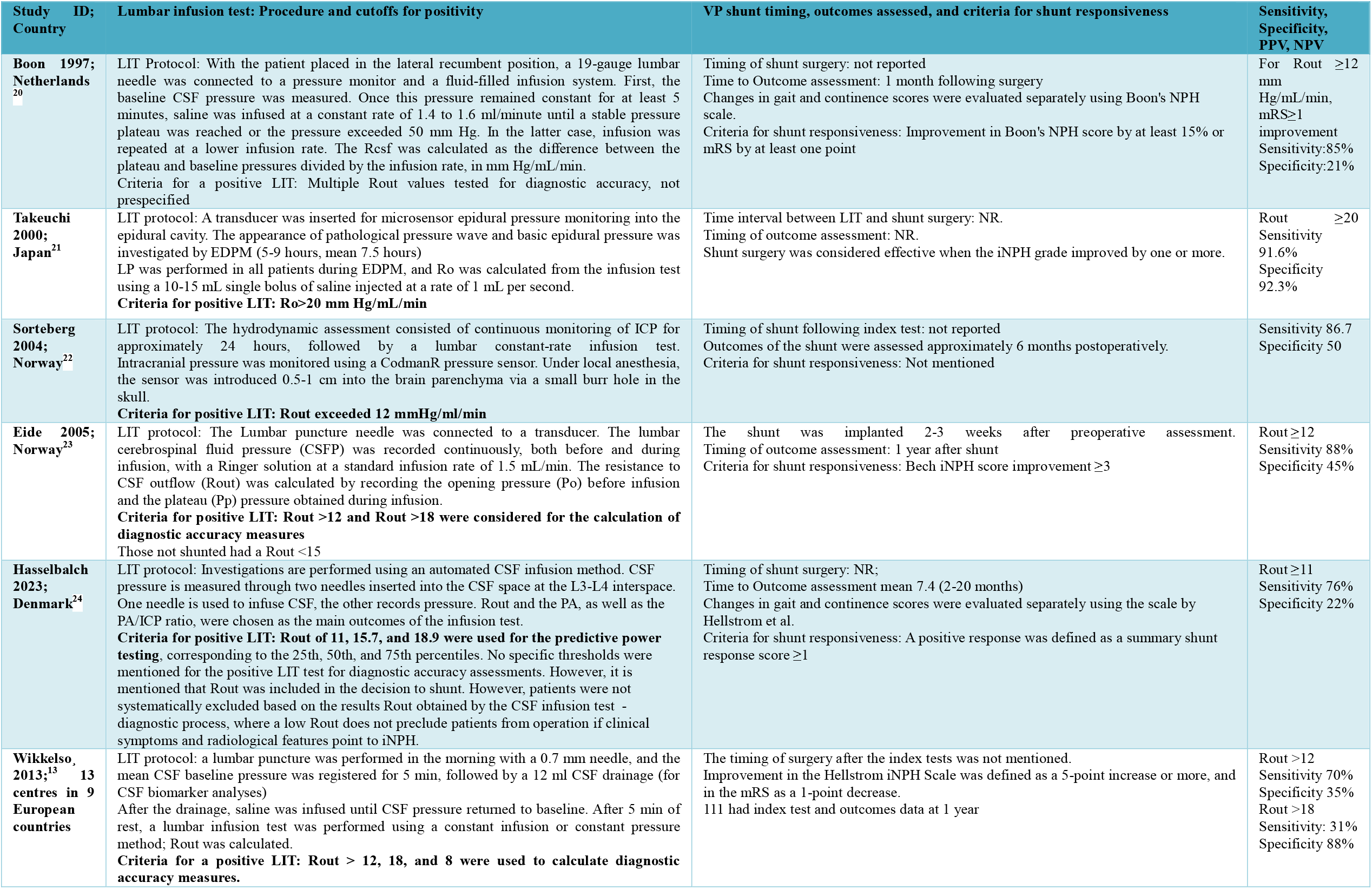
Lumbar Infusion Test: Technique, Assessment, Shunt responsiveness, and diagnostic accuracy.

| Study ID; Country | Lumbar infusion test: Procedure and cutoffs for positivity | VP shunt timing, outcomes assessed, and criteria for shunt responsiveness | Sensitivity, Specificity, PPV, NPV |
| --- | --- | --- | --- |
| Boon 1997; Netherlands<sup>20</sup> | LIT Protocol: With the patient placed in the lateral recumbent position, a 19-gauge lumbar needle was connected to a pressure monitor and a fluid-filled infusion system. First, the baseline CSF pressure was measured. Once this pressure remained constant for at least 5 minutes, saline was infused at a constant rate of 1.4 to 1.6 ml/minute until a stable pressure plateau was reached or the pressure exceeded 50 mm Hg. In the latter case, infusion was repeated at a lower infusion rate. The Rcsf was calculated as the difference between the plateau and baseline pressures divided by the infusion rate, in mm Hg/mL/min. Criteria for a positive LIT: Multiple Rout values tested for diagnostic accuracy, not prespecified | Timing of shunt surgery: not reported<br>Time to Outcome assessment: 1 month following surgery<br>Changes in gait and continence scores were evaluated separately using Boon's NPH scale.<br>Criteria for shunt responsiveness: Improvement in Boon's NPH score by at least 15% or mRS by at least one point | For Rout $\geq 12$ mm Hg/mL/min, mRS $\geq 1$ improvement<br>Sensitivity: 85%<br>Specificity: 21% |
| Takeuchi 2000; Japan<sup>21</sup> | LIT protocol: A transducer was inserted for microsensor epidural pressure monitoring into the epidural cavity. The appearance of pathological pressure wave and basic epidural pressure was investigated by EDPM (5-9 hours, mean 7.5 hours)<br>LP was performed in all patients during EDPM, and Ro was calculated from the infusion test using a 10-15 mL single bolus of saline injected at a rate of 1 mL per second.<br>**Criteria for positive LIT: Ro > 20 mm Hg/mL/min** | Time interval between LIT and shunt surgery: NR.<br>Timing of outcome assessment: NR.<br>Shunt surgery was considered effective when the iNPH grade improved by one or more. | Rout $\geq 20$<br>Sensitivity 91.6%<br>Specificity 92.3% |
| Sorteberg 2004; Norway<sup>22</sup> | LIT protocol: The hydrodynamic assessment consisted of continuous monitoring of ICP for approximately 24 hours, followed by a lumbar constant-rate infusion test. Intracranial pressure was monitored using a CodmanR pressure sensor. Under local anesthesia, the sensor was introduced 0.5-1 cm into the brain parenchyma via a small burr hole in the skull.<br>**Criteria for positive LIT: Rout exceeded 12 mmHg/mL/min** | Timing of shunt following index test: not reported<br>Outcomes of the shunt were assessed approximately 6 months postoperatively.<br>Criteria for shunt responsiveness: Not mentioned | Sensitivity 86.7<br>Specificity 50 |
| Eide 2005; Norway<sup>23</sup> | LIT protocol: The Lumbar puncture needle was connected to a transducer. The lumbar cerebrospinal fluid pressure (CSFP) was recorded continuously, both before and during infusion, with a Ringer solution at a standard infusion rate of 1.5 mL/min. The resistance to CSF outflow (Rout) was calculated by recording the opening pressure (Po) before infusion and the plateau (Pp) pressure obtained during infusion.<br>**Criteria for positive LIT: Rout > 12 and Rout > 18 were considered for the calculation of diagnostic accuracy measures**<br>Those not shunted had a Rout < 15 | The shunt was implanted 2-3 weeks after preoperative assessment.<br>Timing of outcome assessment: 1 year after shunt<br>Criteria for shunt responsiveness: Bech iNPH score improvement $\geq 3$ | Rout $\geq 12$<br>Sensitivity 88%<br>Specificity 45% |
| Hasselbalch 2023; Denmark<sup>24</sup> | LIT protocol: Investigations are performed using an automated CSF infusion method. CSF pressure is measured through two needles inserted into the CSF space at the L3-L4 interspace. One needle is used to infuse CSF, the other records pressure. Rout and the PA, as well as the PA/ICP ratio, were chosen as the main outcomes of the infusion test.<br>**Criteria for positive LIT: Rout of 11, 15.7, and 18.9 were used for the predictive power testing**, corresponding to the 25th, 50th, and 75th percentiles. No specific thresholds were mentioned for the positive LIT test for diagnostic accuracy assessments. However, it is mentioned that Rout was included in the decision to shunt. However, patients were not systematically excluded based on the results Rout obtained by the CSF infusion test - diagnostic process, where a low Rout does not preclude patients from operation if clinical symptoms and radiological features point to iNPH. | Timing of shunt surgery: NR;<br>Time to Outcome assessment mean 7.4 (2-20 months)<br>Changes in gait and continence scores were evaluated separately using the scale by Hellstrom et al.<br>Criteria for shunt responsiveness: A positive response was defined as a summary shunt response score $\geq 1$ | Rout $\geq 11$<br>Sensitivity 76%<br>Specificity 22% |
| Wikkelsø, 2013;<sup>13</sup> 13 centres in 9 European countries | LIT protocol: a lumbar puncture was performed in the morning with a 0.7 mm needle, and the mean CSF baseline pressure was registered for 5 min, followed by a 12 mL CSF drainage (for CSF biomarker analyses)<br>After the drainage, saline was infused until CSF pressure returned to baseline. After 5 min of rest, a lumbar infusion test was performed using a constant infusion or constant pressure method; Rout was calculated.<br>**Criteria for a positive LIT: Rout > 12, 18, and 8 were used to calculate diagnostic accuracy measures.** | The timing of surgery after the index tests was not mentioned.<br>Improvement in the Hellstrom iNPH Scale was defined as a 5-point increase or more, and in the mRS as a 1-point decrease.<br>111 had index test and outcomes data at 1 year | Rout > 12<br>Sensitivity 70%<br>Specificity 35%<br>Rout > 18<br>Sensitivity: 31%<br>Specificity 88% |

Takeuchi et al.^21^ employed epidural pressure monitoring with a 10-15 mL bolus saline injection to calculate Resistance to Outflow (R_out_). Sorteberg et al.^22^ involved prolonged (24-hour) continuous intracranial pressure (ICP) monitoring, followed by a constant-rate infusion test. Eide et al. used a standard infusion rate of 1.5 mL/min of Ringer’s solution, calculating the R_out_ from the opening pressure and plateau pressure. Hasselbalch et al. utilized an automated CELDA System (Likvor AB, Umeå, Sweden) with two needles for infusion and pressure recording, focusing on R_out_, Pulse Amplitude (PA), and PA/ICP ratio. Wikkelsö et al.^13^ performed a baseline pressure measurement, CSF drainage, and saline infusion to return to baseline, followed by a constant infusion or constant pressure method for R_out_ calculation. R_out_ cutoffs selected also differed significantly. Boon et al.^20^ reported the diagnostic accuracy measures at various cutoffs. Takeuchi et al. used >20 mmHg/mL/min^21^; Sorteberg et al.^22^ Eide et al.^23^,, and Wikkelso et al.^13^ used a a cutoff >12 mmHg/ml/min. Wikkelsö et al.^13^ also explored R_out_ >18 and >8. Hasselbalch et al.^24^ used R_out_ percentiles (11, 15.7, 18.9) for predictive power testing without a single fixed threshold for positivity. The presence of pathological pressure waves (Takeuchi et al.^21^) and other parameters, such as PA and the PA/ICP ratio (Hasselbalch et al.^24^), was also considered.

Eide et al.^23^ performed the shunt surgery 2-3 weeks after the LIT, and the other five studies did not report the time interval between the index test and the reference standard. Similar to the studies on the CSF-TT, the criteria for improvement following shunt surgery varied in five studies. Sorteberg et al. reported the number of patients who improved following surgery according to some criteria, which were not explicitly mentioned.^22^ The timing of outcome assessments had similar shortcomings.

The pooled sensitivity of LIT in predicting clinical improvement following shunt surgery was 80.9% (70.3-88.3) with a low between-study variance and heterogeneity (τ^2^ = 0.13, I^2^ 27.6%), and specificity was 42.8% (20.8-68.1) with substantial and heterogeneity (τ^2^ = 1.21, I^2^ 60.8%)(Figure 3) as also depicted in the sROC curve (Supplementary Figure 2) The correlation between sensitivity and specificity was 0.69, implying a positive correlation between sensitivity and specificity across studies. This could reflect low between-study variance.

**Figure 3.**
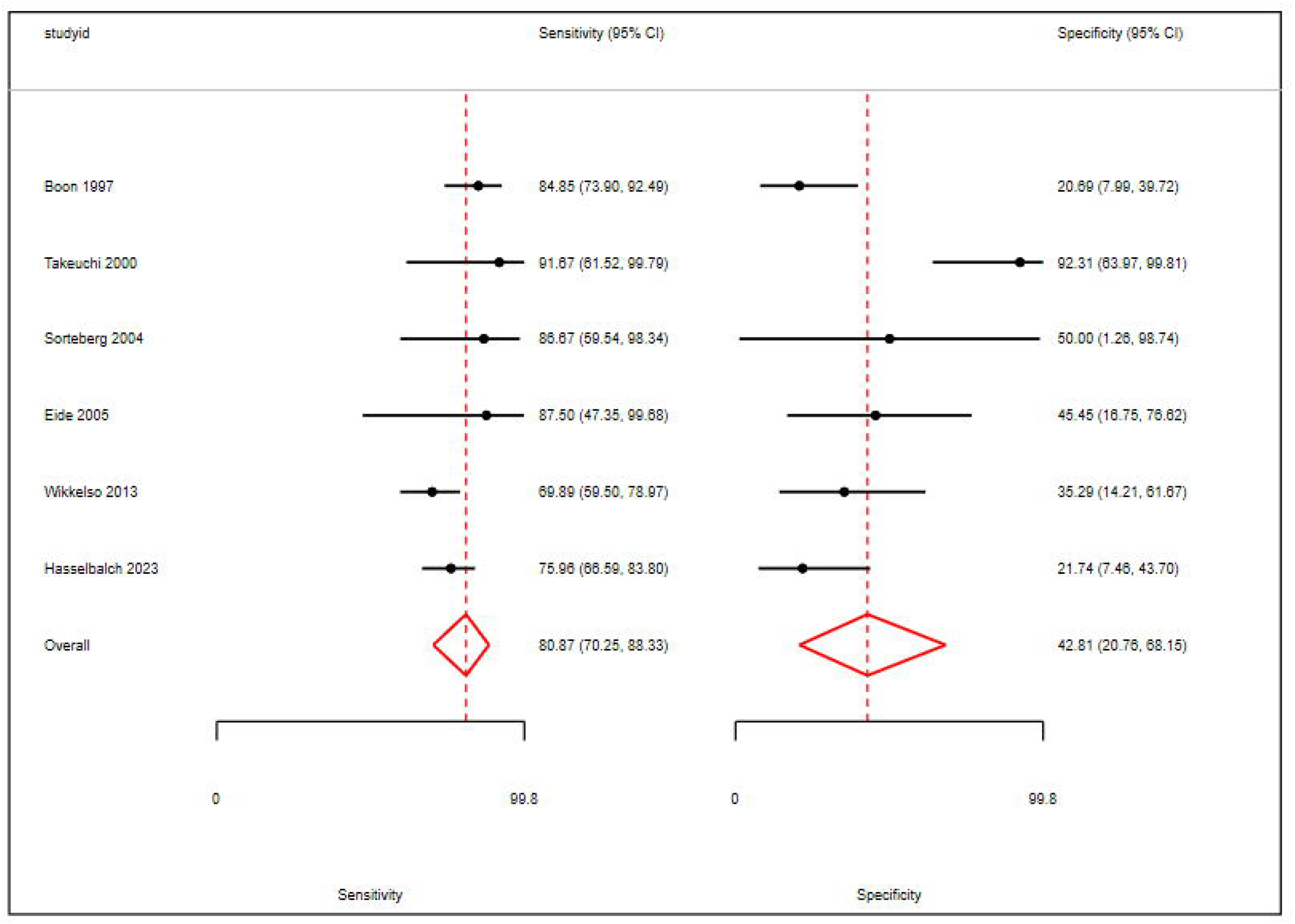
Forest plot showing pooled sensitivity and specificity of the lumbar infusion test (LIT)

### Sensitivity analysis

Since Takeuchi et al. used a cutoff of Rout ≥20, we performed a sensitivity analysis by excluding this study, as the rest of the studies used a cutoff of ≥12, and Hasselbalch et al. used a cutoff of ≥11. The pooled sensitivity was 77.5% (95% CI 70.2-83.4) with very low heterogeneity (τ^2^ = 0.05 and I^2^ = 14.7%) and specificity was 28.3% (95% CI 19-39.9) with very low heterogeneity (τ^2^ = 0.03 and I^2^ = 4.6%). The sensitivity analysis also showed a correlation of -1, suggesting threshold effects.

### Risk of bias

The studies demonstrated various sources of bias, particularly in patient selection, the conduct and reporting of the index tests and reference standards, and in patient flow, as detailed in Table 3 and Supplementary Table 3. Although none of the studies employed a case-control methodology, and most used consecutive series of patients, seven studies had a high risk of bias, and two studies had an unclear risk of bias due to inappropriate patient exclusion, raising concerns about the applicability of the results. Applicability concerns were also identified in several studies, notably Takeuchi et al.^21^, which included only patients with atypical iNPH, Wikkelsö et al.^11^ included patients with secondary NPH (post-subarachnoid hemorrhage, head trauma), and Hasselbalch et al.^24^ had unclear applicability concerns because it was unclear if all clinico-radiologically diagnosed patients were selected for study enrollment.

**Table 3:**
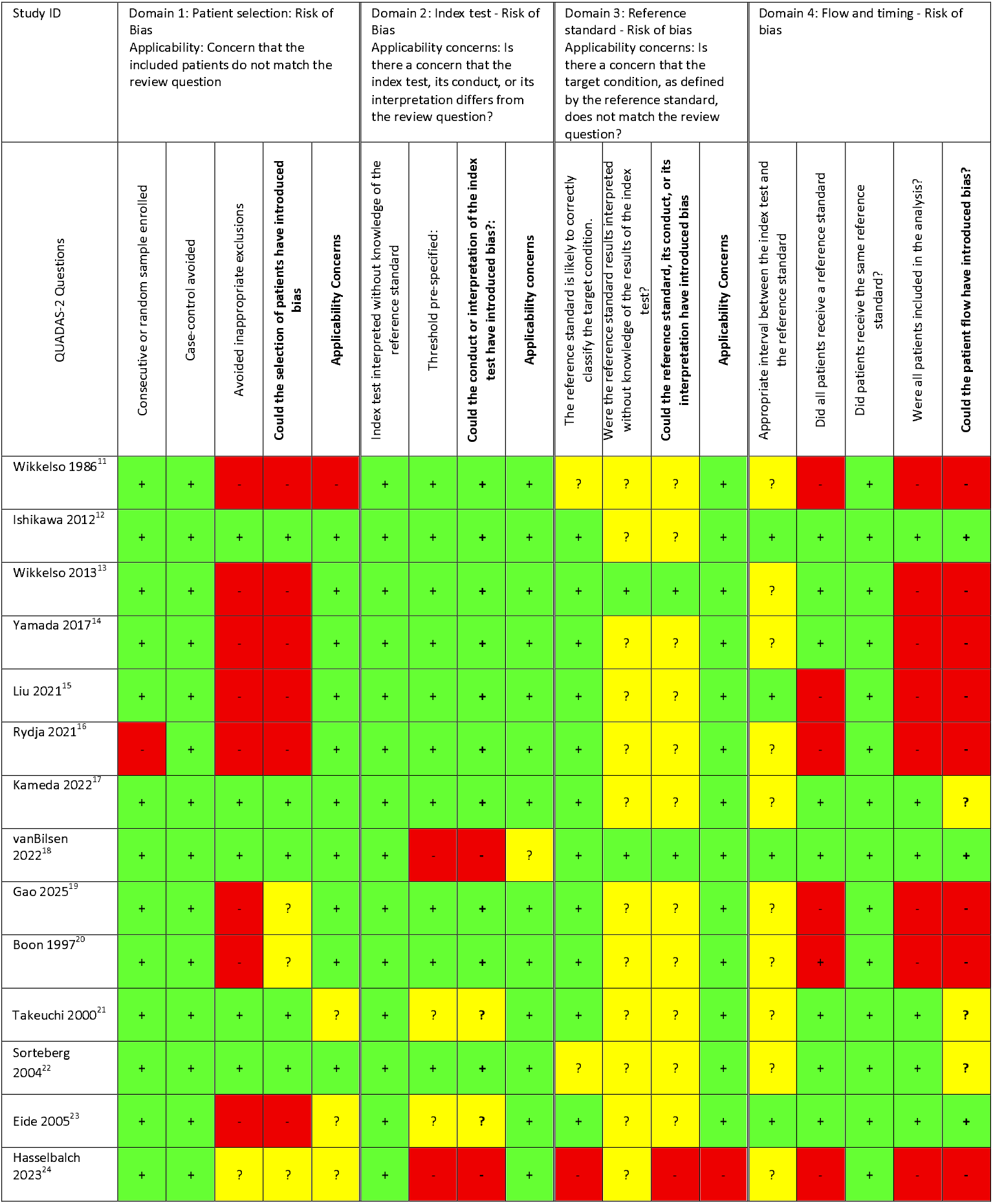
Risk of bias summary: QUADAS 2.

With the index tests, namely CSF-TT and LIT, most studies were reported without knowledge of the reference standard; however, some studies did not report or have pre-specified thresholds, leading to an unknown or high risk of bias. For the reference standard, some studies did not report the criteria for assessing improvement following shunt surgery, thereby making it difficult to determine if the target condition, in this case, iNPH, was correctly identified. Most studies did not report whether the reference standard was interpreted without knowledge of the index test, leading to an unclear risk of bias.

Under the domain flow and timing, only Eide et al.^23^, Ishikawa et al.^12^, and van Bilsen et al.^18^met the criteria for a low risk of bias. Eight studies had a high risk of bias as they did not include all the patients in the analysis (Table 3).

### Summary of Certainty of Evidence

#### CSF Tap Test

The certainty of evidence was very low for sensitivity, downgraded for risk of bias, inconsistency, and imprecision. Based on low-certainty evidence, downgraded for bias and inconsistency, the CSF tap test shows a pooled specificity below 70% (95% CI entirely below 70%) for predicting response to shunt surgery, suggesting that a considerable proportion of non-responders may be incorrectly classified as likely responders. (Supplementary Table 4)

#### Lumbar Infusion Test

The certainty of evidence was moderate for the sensitivity of the LIT to predict response to shunt surgery, downgraded due to serious risk of bias. Additionally, there is moderate certainty evidence, downgraded for bias, the LIT shows a poor specificity (95% CI entirely below 70%) for predicting response to shunt surgery, suggesting that a considerable proportion of non-responders may be incorrectly classified as likely responders. (Supplementary Table 5)

## Discussion

Both the CSF-TT and LIT are widely used diagnostic tools in the evaluation of iNPH. However, there is significant heterogeneity in the protocol for performing these tests, as identified by our systematic review. We also found significant clinical heterogeneity in the timing of assessment following the CSF-TT, type of gait, urinary, or cognitive assessment tools used, cutoffs, and criteria used to assess CSF-TT responsiveness or LIT positivity. Similarly, we found significant differences in the timing of shunt surgery after the index test, the timing of postoperative outcome following the shunt surgery, and the criteria used to define shunt responsiveness. Several studies made no explicit mention of these case definitions, making it difficult, if not impossible, to determine how the diagnosis was established.

The diagnosis of ‘definite’ NPH is made based on response to shunt surgery.^3^ This is somewhat counterintuitive, as the diagnosis, according to this definition, would be made after the treatment. In medicine, we prefer to arrive at a diagnosis prior to offering an invasive surgical procedure as therapy. However, the lack of standardization of diagnostic tests currently impedes the assessment of diagnostic accuracy in NPH.

CSF-TT is generally considered less invasive and simpler to perform. It also gives clinicians a sense that, since it mimics CSF drainage by shunt surgery, it intuitively makes sense that only patients with a positive tap test should undergo definitive surgery, thereby making it a common initial screening tool for referring patients for surgical treatment. However, the currently available literature does not support this notion. The guidelines, therefore, state that, regardless of the tap test results, patients may be referred for surgery, as is the practice in Japan. In line with these recommendations, we found that the sensitivity of the CSF-TT was 67.5% (95% CI 52-80%), meaning that several patients who were CSF-TT non-responders still responded to VP shunting.

In contrast, although the LIT is a more invasive procedure, it is considered to provide quantitative physiological data on CSF dynamics, such as R_out_ and ICP PA. While some studies report exceptionally high accuracy (e.g., Takeuchi et al,^21^ reported 91.6% sensitivity and 92.3% specificity using a R_out_ cutoff ≥20 mmHg/ml/min), others show concerningly low specificity (e.g., 22% specificity with a cutoff ≥11 in Hasselbalch et al,^24^), indicating a potential risk of false positives. The lack of consensus on pathological R_out_ values and the influence of methodological variations on measured parameters remain significant limitations. These variations in infusion methods, pressure monitoring techniques, and the specific parameters measured make it difficult to compare these studies uniformly.

Both the CSF-TT and LIT were evaluated using the GRADE approach to assess the certainty of evidence for diagnostic accuracy in predicting response to shunt surgery in patients with probable iNPH. The CSF-TT demonstrated a sensitivity of 67.5% (95% CI: 52–80); however, the certainty of evidence was rated very low. This was due to a serious risk of bias in the included studies, a very serious inconsistency arising from statistical heterogeneity (evidenced by a high τ^2^ and I^2^), and serious imprecision, as the lower bound of the confidence interval crossed the threshold of diagnostic utility of 70%. In contrast, there is low-certainty evidence that the specificity of CSF-TT was poor at 53% (95% CI: 40.7–65.5), due to a serious risk of bias and serious inconsistency.

Similarly, the LIT showed a high pooled sensitivity of 80.9% (95% CI: 70.3-88.3). The certainty of evidence was rated as moderate, downgraded due to a serious risk of bias resulting from methodological limitations across the studies. The specificity of LIT was poor at 42.8% (95% CI: 20.8-68.1), with moderate certainty of evidence, downgraded due to a serious risk of bias. We did not downgrade for inconsistency because Takeuchi et al. used a different cutoff, and the sensitivity analysis excluding this study revealed no significant heterogeneity.

These findings underscore that while CSF-TT and LIT may help identify patients who could benefit from shunt surgery, their limited specificity and very low certainty of evidence caution against their use as standalone diagnostic tools. There remains a critical need for higher-quality studies with standardized protocols and blinded assessments to better define their diagnostic value. The inconsistent performance of both tests, particularly when relying on clinical outcome measures or fixed cutoffs, strongly suggests that NPH shunt responsiveness is a complex, multifactorial phenomenon that is unlikely to be captured by a single, simple test. If multiple underlying pathophysiological mechanisms influence a complex medical condition, it is highly improbable that a single diagnostic marker will be perfectly predictive of treatment response. This reinforces the need for a standardized multi-modal assessment approach that integrates various clinical, radiological, and physiological parameters.

Our review has a few limitations that warrant consideration. We did not study ELD, which is used in several settings, albeit infrequently. Many of the included studies were retrospective in design and involved heterogeneous patient populations. In some cases, outcome reporting was incomplete, and patients with surgical complications — such as postoperative bleeding — were excluded from final analyses, which may influence the interpretation of results. Additionally, there was variation in the protocols used for index tests and outcome assessment, reflecting the lack of standardized approaches at the time. Several of these studies were conducted decades ago when reporting standards were still evolving, and this is reflected in the overall methodological quality. As a result, concerns regarding the risk of bias and applicability were raised in several studies. Therefore, the findings on diagnostic accuracy should be interpreted with these limitations in mind. We did not perform a sensitivity analysis because there were very few studies without a significant risk of bias, and thus, it would not make sense to exclude those with a high risk of bias. Heterogeneity of studies is another major issue. Pooled estimates are derived from highly heterogeneous protocols. Thresholds for test positivity, timing of outcome assessment, and definition of ‘shunt response’ varied significantly, limiting the external validity and generalizability of the results. We did not assess for publication bias, and funnel plots were not generated due to the limited number of studies per test; therefore, small-study effects could not be ruled out.

## Conclusion

The role of the CSF-TT and LIT in predicting outcomes following shunt surgery for NPH appears to be modest at best. In our analysis, both tests demonstrated low pooled specificity — 53.3% for CSF-TT and 42.8% for the LIT, indicating a limited ability to accurately distinguish between true non-responders and those who may benefit from surgery as standalone tools. In practical terms, this level of diagnostic performance of the CSF-TT is comparable to the randomness of a coin toss, raising concerns about the reliability of the test when used in isolation to guide clinical decisions. On the other hand, there is moderate-quality evidence to suggest that the LIT has poor specificity, although the included studies suffered from a significant risk of bias.

Given these findings, clinicians should exercise caution when using the results of these tests to prognosticate surgical outcomes. Relying solely on the CSF-TT may lead to both over- and under-treatment, especially since only half of patients with positive test results have good outcomes following surgery, and up to 32% of those with negative test results have favorable outcomes. The test results of the tap test should not be interpreted in isolation, and management paradigms should involve looking holistically at the clinical picture, imaging, and also the patients’ values and preferences, given that current tests are not sufficiently valid to predict postoperative outcomes. Prospective studies using standardized protocols with clearly defined outcome parameters following shunt surgery are needed.

Moving forward, there is a clear need to develop and validate more accurate and comprehensive predictive models that integrate clinical, radiological, and perhaps biomarker data. Machine learning models incorporating clinical, imaging, and biomarker data should be explored. These will ensure better predictive models for post-shunt recovery in iNPH. Such models could not only improve patient selection for shunt surgery but also enhance the process of shared decision-making by providing patients and families with clearer, evidence-based expectations regarding treatment outcomes.

## Data Availability

none generated

## Legends

Supplementary Figure 1. Summary receiver operating characteristic (SROC) curve for CSF-TT

Supplementary Figure 2. Summary receiver operating characteristic (SROC) curve for LIT

Supplementary Table 1. Characteristics of included studies reporting the diagnostic accuracy of the CSF-TT

Supplementary Table 2. Characteristics of included studies reporting the diagnostic accuracy of the LIT

Supplementary Table 3. Methodological quality assessment using QUADAS-2

Supplementary Table 4. GRADE summary of findings table for the CSF-TT

Supplementary Table 5. GRADE summary of findings table for the LIT

Appendix 1: Search strategy

## Funding

No funding received

Patients or the public WERE NOT involved in the design, conduct, reporting, or dissemination plans of our research.

