## Supplementary Table for "Accuracy of CSF Tap Test and Lumbar Infusion Test in Predicting Shunt Response in Idiopathic Normal Pressure Hydrocephalus: A Systematic Review and Meta-Analysis"

### Supplementary Table 1: Demographics and Details of studies on CST-TT

| **Study ID; Country; Duration**  **Aims, Population description, Inclusion and exclusion criteria** | **Mean age, Clinical profile, Evans index, and baseline scores** |
| --- | --- |
| 1. **Wikkelso 1986**;^1^ Sweden; 1979-1983; Diagnostic test accuracy study   Funding: grants from the Edit Jacohson's Foundation, the Medical Faculty of Gothenburg, the Medical Society of Gothenburg, and the Swedish Medical Research Council  Aim of the study: Evaluation of the predictive value of the CSF-TT  Participants: Patients who were investigated for NPH and underwent VP shunt  Inclusions: consecutive patients evaluated and operated with VP shunt; only 6 had iNPH, 9 had SAH, 8 had head trauma, and 4 had cerebrovascular diseases  Selection for shunt surgery was based on characteristic changes observed during clinical investigation, CT, and radionuclide cisternography. Inflammatory and metabolic disorders were excluded by appropriate CSF and blood examinations; 3 patients with complications were excluded from the analysis.  **CSF Tap test was not used in the decision-making process for shunting** | **27 patients**; Mean age: 63 (40-81); M/F: 13/14;  Vascular disease: NR; Cognitive impairment: NR  Gait dysfunction: 89%, Urinary dysfunction: 81%; Duration of symptoms: 21 months (3 months – 9 years)  Evans Index: 0.38 (0.30-0.46)  Baseline Gait score, cognitive and urinary scores: NR  CSF opening pressure: 12±5mm Hg  **Index test and outcome data available in 21 patients** |
| 2. **Ishikawa 2012**;^2^ Japan; 2004-2006; Prospective cohort study   Funding: Supported in part by Johnson and Johnson, Nihon Medi Physics, and Daichi Pharmaceuticals, honoraria from companies that manufactured devices;  Aim of the study: The predictive value of TT was investigated in patients with iNPH; validate the diagnostic importance of high-convexity tightness in coronal-section MRI with the results of shunt surgery using a programmable valve  Participants: Patients with iNPH who underwent VP shunt  Inclusions: (1) 60 to 85 years old, (2) one or more of the NPH triad symptoms, (3)Ventriculomegaly (Evans Index > 0.3), (4) high-convexity tightness in coronal-section MRI, and (5) no antecedent disorders.  **CSF Tap test was not used in the decision-making process for shunting** | **100 patients**; 80 shunt responders, 20 non-responders;  Median age: 75; M/F: 58/42  Vascular disease: na; Cognitive impairment: 80%; Gait dysfunction: 91%; Urinary dysfunction: 60%  Duration of symptoms: NR; Evans Index: 34.6 (32.7-38); Baseline iNPH Gait score: 2 (2-3); Baseline iNPH Cognitive score: 2 (1-3); Baseline urinary score: 2 (1-3.7) Mean volume of CSF tapped: 30 mL; CSF opening pressure: 12(9-14) cm H20  **Index test and outcome data available in 100 patients** |
| 3. **Wikkelso¸ 2013**;^3^ 13 centers in 9 European countries; Sep 2004-Jan 2008; Prospective cohort study   Funding: Codman & Shurtleff, Raynham, Massachusetts, USA  Aim of the study: To determine the sensitivity, specificity, and positive and negative predictive values of the CSF TT and Rout in patients clinically presenting with iNPH.  Participants: iNPH; Inclusions: Gradually developed gait disturbance affecting tandem walking, turning, stride length, and width. The MRI criteria for iNPHT were a symmetrical quadri-ventricular enlargement without cortical infarcts or other clinically relevant parenchymal lesions (lacunar infarcts of less than 1 ml were accepted), whereas single cortical infarcts were accepted for a diagnosis of iNPHQ. Free communication between the ventricular system and the subarachnoid space and an Evans Index greater than 0.30 were mandatory for both diagnoses.  Exclusions: Patients who were unwilling to participate, had a restricted life expectancy, or had contraindications to surgery  **CSF Tap test was not used in the decision-making process for shunting** | **115 patients**, Mean age: 71(9); M/F: 58/57; Vascular disease: 8%; Cognitive impairment, Urinary and gait dysfunction: Not reported  Duration of symptoms: 18 months (12-30)  Evans Index: 0.42 (0.31-0.66); Baseline Gait score, cognitive and urinary scores: NR  Mean volume of CSF tapped: 50ml; CSF opening pressure: 10 mmHg(2-18)  **111 had index test and outcome data at 1 year** |
| 4. **Yamada 2017**;^4^ Japan; March 2010-Oct 2011; Cohort study   Funding: The funding sources (Johnson & Johnson K.K. and NihonMedi-Physics Co. Ltd.)  Aim of the study: Investigate time duration since the initial presentation of iNPH symptoms until the CSF tap test as a pivotal factor that could increase the accuracy of predicting the iNPH diagnosis using the CSF tap test.  Participants: Possible iNPH according to the second edition of the Japanese iNPH guideline  Inclusions: Age at entry 60-85 years; patients with one or more of gait disturbance, cognitive impairment, urinary disturbance on the iNPH grading scale (iNPHGS) within 3 mo before date of consent; ventriculomegaly with Evans index >0.3, in concurrent with narrow sulci at the high convexity and enlarged Sylvian fissure on CT or MRI.  Exclusions: Those with a diagnosis of secondary hydrocephalus that occurred after subarachnoid hemorrhage, meningitis, head trauma, congenital hydrocephalus, or aqueductal stenosis, among others; CSF pressure of ≥20 cm H2O; complications of severe disuse muscle atrophy; psychiatric disorders or other neurological diseases  **CSF Tap test was not used in the decision-making process for shunting** | **93 patients**; Mean age (SD) 76.5(4.5)  Vascular disease: NR Cognitive impairment, Gait dysfunction, urinary dysfunction: NR  Duration of symptoms: 24.2(1-120) mo Evans Index: NR  Baseline iNPH Gait score 2.3-2.5(0.7) iNPH Cognitive score 2.3-2.4 (0.9) and urinary score 2 (0.9) iNPH total score 6.6 (2.2)  Mean volume of CSF tapped, CSF opening pressure: NR  **82 patients had index tests and reference standard outcomes available.** |
| 5. **Liu 2021**;^5^ China; 2012-2019; Retrospective cohort study   Funding: Grant from National Key Research and Development Program of China, CAMS Innovation Fund for Medical Sciences (2016-12M-1-004), National Natural Science Foundation of China, and the Strategic Priority Research Program Biological basis of aging and therapeutic strategies of the Chinese Academy of Sciences  Aim of the study: To compare the improvement in TUG, 10 m walking time, 10 m walking steps, and composite Z score at 8 h, 24 h, and 72 h after CSF TT and calculate the specificity, sensitivity, PPV and NPV of the walking tests and executive composite Z score according to the results of shunting follow-up assessments  Participants: clinically possible normal-pressure hydrocephalus according to international diagnostic criteria and Chinese consensus  Inclusions: 1. diagnosis with iNPH according to international guidelines and Chinese consensus; 2. completion of a multi-time point assessment of walking ability and neuropsychological status.; Exclusions: The patients could not tolerate 30 ml of CSF drainage during the CSF TT  **CSF Tap test was not used in the decision-making process for shunting** | **88 patients**; Age 67±12 y  M/F: 61/27; Vascular disease: NR  Cognitive impairment: 71.6%; Gait impairment: 84%; urinary dysfunction: 67%  Duration of symptoms: 0.5-11 years  Evans Index, Baseline Gait, urinary and cognitive scores: NR  **Index test and outcomes of reference standards available in 23 patients** |
| 6. **Rydja 2021**;^6^ Sweden; Jan 2016-Dec 2019; Retrospective cohort study   Funding: Open access funding provided by Linkoping University  Aim of the study: To evaluate the prognostic value of the CSF TT using the Hellstrom iNPH scale among shunted iNPH patients in a large single-center cohort  Participants: Patients with a diagnosis of possible and probable iNPH, based on the international guidelines of 2005 and  treated with an adjustable valve shunt  Inclusions: All patients had a disturbed gait and balance and, additionally, an impairment of cognition and/or continence symptoms. Exclusions: Patients were excluded if they had missing data for the CSF TT or the post-operative assessment.  **CSF Tap test was not used in the decision-making process for shunting** | **159 Patients**; Age: 75.4 (6.2); M/F: 62/54;  Vascular disease: 9.5% Cognitive impairment, Gait dysfunction, and urinary dysfunction: NR  Duration of symptoms, Evans Index, Baseline Gait, and urinary scores: NR  Baseline Cognitive score: MMSE 26 (23-28) n=115; Total iNPH score 51.8(16.9)  Mean volume of CSF tapped: 48.0 (44.0-50.0)  CSF opening pressure: 15 (12.5-18.0) cm H20  **Index test and outcomes of reference standard available in 116 patients.** |
| 7. **Kameda 2022**;^7^ Japan; Apr 2021-Sep 2021; Retrospective cohort study   Funding: Japan Society for the Promotion of Science Grant-in-Aid for Scientific Research;  Aim of the study: To study if the comprehensive evaluation of the effect of the tap test using the Functional Gait Assessment (FGA) and Global Rating of Change (GRC) scales can lead to more appropriate patient selection for shunting than the evaluation of the effects of TUG alone  Participants: Patients with a positive tap test who showed some improvement in any of the three symptoms—gait, urinary incontinence, and cognitive function — after the tap test were determined to be eligible for LP shunt. In addition, patients with a negative tap test but DESH findings were identified as having a high probability of iNPH and were considered eligible for LP shunt.  **CSF Tap test was not used in the decision-making process for shunting** | **40 patients;** Mean age: 78.8(5.2)  M/F: 19/21  Vascular disease, Cognitive impairment, Gait dysfunction, urinary dysfunction: NR  Duration of symptoms, Evans Index: NR  Baseline TUG 15.3 (5.7); MMSE 24.5 (4.6); iNPH 5.2(2.4)  **TUG test outcomes available in 40, FGA in 24 patients** |
| 8. **vanBilsen 2022**;^8^ Netherlands; Jan 2015-Jan 2020; Prospective cohort study   Funding: Not reported  Aim of the study: A high delta amplitude would be a good diagnostic marker for a positive shunt response, and a low amplitude or inability to lower the amplitude would correlate with a negative shunt response. Combining delta amplitude with other diagnostic markers deduced from LIT, such as P0, Rout, and tap test, could improve specificity and sensitivity.  Participants: iNPH; Inclusions: Patients who underwent shunt surgery preceded by a LIT according to the new pulsatility curve protocol for diagnosis of iNPH  Exclusions: secondary NPH or aqueductal stenosis  **CSF Tap test was not used in the decision-making process for shunting** | 38 patients; Mean age: 72; M/F: 24/14;  Vascular disease: 37%; Cognitive impairment: 32(84.2); Gait dysfunction: 100%, urinary dysfunction: 82%  Duration of symptoms, Evans Index, Baseline Gait, Cognitive and urinary scores: NR  Pre-op iNPH score (median) 5.5 (5-7)  **Index test and outcomes available in 38 patients** |
| 9. **Gao 2025,**^9^ China, Aug 2019-Nov 2023; Retrospective cohort study   Funding: Key R&D Program of Zhejiang Province, National Natural Science Foundation of China, A Project Supported by Scientific Research  Fund of Zhejiang University  Aim of the study: To study the predictive performance of preoperative imaging features combined with tap test for the outcomes of ventriculoperitoneal (VP) shunt in idiopathic normal pressure hydrocephalus (iNPH).  Participants: diagnostic criteria were based on the second edition of the Japanese iNPH Treatment Guidelines  Exclusions: All possible causes of secondary hydrocephalus,Blurred magnetic resonance imaging (MRI) images  **CSF tap test was not used in the decision making process for shunting** | 211 patients; Mean age: 71(7); M/F: 110/56  Vascular disease: 44%; Cognitive impairment: 77%; Gait dysfunction: 97%; urinary dysfunction: 79%  Duration of symptoms: 19(7.4 months)  Evans index: 0.35  Baseline scores: Not reported  CSF opening pressure: 12.9 cm in responder 13.4 in non responders  Index test and outcomes available in 166 patients |

### Supplementary Table 2: Demographics and Details of studies on LIT

| Study ID; Country; Duration  Aims, Population description, Inclusion and exclusion criteria | Number of patients; Mean age; Clinical features, Evans index, baseline gait, urinary and cognitive scores | Plateau pressure, resistance to outflow, pulse pressure amplitude, baseline ICP |
| --- | --- | --- |
| Boon 1997; Netherlands;^10^ September 1990-July 1995; Prospective cohort study  Funding: Doctor Ed. Hoelen Stichting and a foundation that wishes to remain anonymous  Aim of the study: To study if the measurement of resistance to outflow of cerebrospinal fluid (Rcsf) predicts outcome following surgery in iNPH  Participants: 1) a gradually developed gait disturbance of both legs, unexplained by other conditions, and a gait scale score of at least 12; 2) a mild-to-moderate cognitive deficit without aphasia, emerging with or after the gait disturbance, and a dementia scale score of at least 12; 3) a disability mRS grade of at least 2; and 4) a computerized tomography (CT) scan showing a communicating hydrocephalus with an Evans’ index of 0.3 or greater and a ventricular index greater than 0.8,4 without clinically relevant parenchymal lesions, the sum of the four largest sulci at the convexity being less than 25 mm  Exclusions: Acute or subacute symptomatic NPH within 3 months of a causative incident, age of 85 years or more, severe comorbidity with restricted life expectancy, or contraindications for surgery | No of patients 101; Mean age: 74(6.3); Range M/F:60/41  Vascular disease; Cognitive impairment, Gait dysfunction, urinary dysfunction - NR  Duration of symptoms: 2.3±2.3 years Evans Index: 0.39±0.06  Baseline Gait score: 25.6±10.2  Baseline Cognitive score: 23.3±7.6; Baseline mRS: 3.7±1.3  Index test and outcomes available in 95 patients | Plateau pressure, Resistance to outflow, Pulse pressure amplitude, PA ICP ratio NR  Baseline ICP 11.2±3.3 |
| Takeuchi 2000; Japan;^11^ July 95-Dec 97: Cohort - not clear if prospective or retrospective  Funding: Japanese Ministry of Health and welfare grant in aid for scientific research into intractable hydrocephalus  Aim of the study: To study the effectiveness of shunt surgery in patients with iNPH with cerebral atrophy (atypical iNPH)  Participants: Idiopathic NPH presenting with Dementia and Brain atrophy; Inclusions: main complaint of dementia, sometimes with gait disturbance; intermediate or severe cerebral atrophy, bilateral symmetrical ventricular dilatation and at least periventricular low density at the anterior horn on CT; and CSF pressure within the normal range with inflows of contrast medium into the cerebral ventricle or cortical surface 24 hours later on cisternography | No of patients 25  Mean age: 60.4 (47-83); M/F: 15/10;  Vascular disease: NR; Cognitive impairment, Gait dysfunction, urinary dysfunction: NR  Duration of symptoms: NR  Evans index and baseline scores: NR  Index test and reference standard outcomes available in 25 | Plateau pressure: NR  Resistance to outflow: responder: 35.33 (11.16) non-responder: 9.12 (3.51)  Pulse pressure amplitude: NR; baseline ICP: NR; PA ICP ratio NR; Pressure wave seen in 66.7% of shunt responders and 0% of shunt non-responders |
| Sorteberg 2004;^12^ Norway; Prospective cohort study  Funding: Not reported  Aim of the study: To study the predictive value of the lumbar infusion test with computation of resistance to cerebrospinal fluid (CSF) outflow (Rout) and continuous ICP monitoring with computation of baseline or mean ICP and frequency of ICP elevations  Participants: Inclusions: Clinical signs of NPH, including gait disorder, urinary incontinence, and decreased memory; radiological findings of ventriculomegaly (Evans index >0.3)  Exclusions: Other diseases causing symptoms indicative of NPH; Non-idiopathic hydrocephalus; incomplete pre-operative work-up. | No of patients 17; Mean age: 65; M/F: 7; Vascular disease: NR ; Cognitive impairment:88.2%  Gait dysfunction: 94%  Urinary dysfunction: 88.2%  Duration of symptoms: 1.5 years (6 months – 6 years; Evans index: >0.3 in all  Index test and reference standard outcomes available in 17 | Plateau pressure: 37.14 +8.13 mmHg  Resistance to outflow: 16.55(4.05)(8.7-23.7) mmHg/ml/min  Baseline ICP: 8.75 ± 3.88 mmHg |
| Eide 2005;^13^ Norway; April 2002-March 2003; Retrospective cohort  Funding: Not reported  Aim of the study: To analyze the pre-operative spinal hydrodynamics during lumbar infusion tests of all iNPH cases treated with shunts and to evaluate to what extent these findings correlated to changes in the NPH score 1 year after shunt treatment.  Participants: Inclusions: Patients with the presence of minimum two deficits (gait disturbance, urinary incontinence, or dementia) and an Evans index > 0.3, treated with shunts | No of patients: 19 Mean age: 68 y (Range 47-81); M/F: 8/11 Vascular disease: 26% Cognitive impairment, Gait dysfunction, Urinary dysfunction: NR Duration of symptoms: 2.5 years (1-35) Evans Index: >0.3 in all; Pre-operative iNPH 9 (5-14), postoperative iNPH 11 (4-15)  Index test and reference standard outcomes available in 19 | Plateau pressure: NR Resistance to outflow: median 16.3 mmHg/ml/min (ranges 4.6-21.0); Pulse pressure amplitude: baseline ICP: PA ICP ratio NR. Mean CSFP wave amplitudes during LIT: significantly higher in shunt responders than in the non-responders. |
| Hasselbalch 2023;^14^ Denmark; 2013-2020; Retrospective cohort  Funding: Danish Dementia Research Centre  Aim of the study: To investigate the predictive power of Rout, PA, and the PA/ICP ratio and to combine the best of the two with the iNPH Radscale in an attempt to increase the ability to predict short-term shunt response in patients with iNPH.  Participants: Patients with probable or possible NPH; Inclusions: (1) diagnosis of probable or possible iNPH according to international guideline criteria (2) lumbar infusion test as part of the diagnostic evaluation (3) an available brain CT or MRI scan, performed within 1 year prior to surgery (4) subsequent ventriculoperitoneal shunt (5) at least 2 months of clinical follow-up after operation | No of patients 127; Mean age: 74±6; Range 55-87 M/F: 46/81  Vascular disease: NR Cognitive impairment, Gait dysfunction, urinary dysfunction - NR  Duration of symptoms: 3.7±3.1 years Evans Index: Not reported  Baseline Gait score: 3.9±1.7(1-8)  Baseline Cognitive score: 25.2±4.1(11-30); Baseline urinary score: 2.9±1.2; (0-6)  Index test and reference standard outcomes available in 127 | Plateau pressure: NR  Resistance to outflow: 15.9±6.5; 3.7-48.5; Pulse pressure amplitude: 3.8±1.9; Baseline ICP: 11.3±3.1; PA/ICP Ratio: 0.34 ± 0.17. |
| Wikkelso¸ 2013;^3^ Details in Supplementary table 1 | Details in Supplementary table 1  Index test and reference standard outcomes available in 110 | Plateau pressure, Resistance to outflow, Pulse pressure amplitude, baseline ICP, PA ICP ratio NR |

### Table 3: Risk of bias assessment QUADAS 2

| Study ID | Domain 1: Patient selection: Risk of Bias  Applicability: Concern that included patients do not match review question | Domain 2: Index test - Risk of Bias  Applicability concerns: Is there concern that the index test, its conduct, or its interpretation differ from the review question? | Domain 3: Reference standard - Risk of bias  Applicability concerns: Is there a concern that the target condition, as defined by the reference standard, does not match the review question? | Domain 4: Flow and timing - Risk of bias |
| --- | --- | --- | --- | --- |
| 1. Wikkelso 1986^1^ | Consecutive or random sample enrolled: Yes  Case-control avoided: Yes  Avoided inappropriate exclusions: No, Only 21 patients underwent evaluation with a walk test, and only 24/27 had post-operative outcomes.  **Could the selection of patients have introduced bias? High**  **Applicability concerns: High** (post-SAH, head trauma included) | Index test interpreted without knowledge of reference standard: Yes  Threshold pre-specified: Yes  **Could the conduct or interpretation of the index test have introduced bias?: Low**  **Applicability concerns: Low** | Reference standard likely to correctly classify the target condition: Unclear (no criteria)  Were the reference standard results interpreted without knowledge of the results of the index test: Unclear.  **Could the reference standard, its conduct, or its interpretation have introduced bias: Unclear**  **Applicability concerns: No** | Appropriate interval between index test and reference standard: Unclear  Did all patients receive a reference standard? No  Did patients receive the same reference standard? Yes  Were all patients included in the analysis? No  **Could the patient flow have introduced bias? High** |
| 2. Ishikawa 2012^2^ | Consecutive or random sample enrolled: Yes Case-control avoided: Yes  Avoided inappropriate exclusions: Yes  **Could the selection of patients have introduced bias: Low**  **Applicability Concerns: Low** | Index test interpreted without knowledge of reference standard: Yes  Threshold pre-specified: Yes.  **Could the conduct or interpretation of the index test have introduced bias: Low**  **Applicability concerns: Low** | Reference standard likely to correctly classify the target condition: Yes  Were the reference standard results interpreted without knowledge of the results of the index test: Unclear  **Could the reference standard, its conduct, or its interpretation have introduced bias: Unclear**  **Applicability concerns: Low** | Appropriate interval between index test and reference standard: Yes (done within 2 months)  Did all patients receive a reference standard? Yes  Did patients receive the same reference standard? Yes  Were all patients included in the analysis? Yes  **Could the patient flow have introduced bias? Low** |
| 3. Wikkelsø 2013^3^ | Consecutive or random sample enrolled: Yes  Case-control avoided: Yes  Avoided inappropriate exclusions: No (115/142 only had outcomes at 1 y)  **Could the selection of patients have introduced bias? High**  **Applicability Concerns: Low** | Index test interpreted without knowledge of reference standard: Yes  Threshold pre-specified: Yes  **Could the conduct or interpretation of the index test have introduced bias: Low**  **Applicability concerns: Low** | Reference standard likely to correctly classify the target condition: Yes  Were the reference standard results interpreted without knowledge of the results of the index test: Yes  **Could the reference standard, its conduct, or its interpretation have introduced bias: Low**  **Applicability concerns: Low** | Appropriate interval between index test and reference standard: Unclear  Did all patients receive a reference standard? Yes  Did patients receive the same reference standard? Yes  Were all patients included in the analysis?No  **Could the patient flow have introduced bias? High** |
| 4. Yamada 2017^4^ | Consecutive or random sample enrolled: Yes  Case-control avoided: Yes  Avoided inappropriate exclusions: No (82/93 had final data available)  **Could the selection of patients have introduced bias? High**  **Applicability Concerns: Low** | Index test interpreted without knowledge of reference standard: Yes  Threshold pre-specified: Yes  **Could the conduct or interpretation of the index test have introduced bias: Low**  **Applicability concerns: Low** | Reference standard likely to correctly classify the target condition: Yes  Were the reference standard results interpreted without knowledge of the results of the index test: Unclear  **Could the reference standard, its conduct, or its interpretation have introduced bias: Unclear**  **Applicability concerns: Low** | Appropriate interval between index test and reference standard: Unclear  Did all patients receive a reference standard? Yes  Did patients receive the same reference standard? Yes  Were all patients included in the analysis?No  **Could the patient flow have introduced bias? High** |
| 5. Liu 2021^5^ | Consecutive or random sample of patients enrolled: Yes  Case-control avoided: Yes  Study avoided inappropriate exclusions: No (only 88 patients underwent index test, and only 23 out of 115 patients admitted for evaluation had outcome data available.  **Could the selection of patients have introduced bias? High**  **Applicability concerns: Low** | Index test interpreted without knowledge of reference standard: Yes  Threshold pre-specified: Yes  **Could the conduct or interpretation of the index test have introduced bias: Low**  **Applicability concerns: Low** | Reference standard likely to correctly classify the target condition: Yes  Were the reference standard results interpreted without knowledge of the results of the index test: Unclear  **Could the reference standard, its conduct, or its interpretation have introduced bias: Unclear**  **Applicability concerns: Low** | Appropriate interval between index test and reference standard: Yes (Done within one month)  Did all patients receive a reference standard?No  Did patients receive the same reference standard? Yes  Were all patients included in the analysis?No  **Could the patient flow have introduced bias? High** |
| 6. Rydja 2021^6^ | Consecutive or random sample of patients enrolled: No; Out of 159 iNPH patients, 116 patients (95 patients with probable iNPH and 21 with possible iNPH) were included in the statistical analysis. 43 were excluded: 25 did not undergo the CSF TT, five had been investigated with external lumbar drainage (ELD), 13 patients had no result from the follow-up post-operative assessment (six had shunt complications, five had missing data due to unwillingness to participate and two died). One of the deaths was caused by an acute subdural hematoma 2 months after shunt surgery, and the other by an intracerebral hematoma 5 days after surgery.  Case-control avoided: Yes.  Avoided inappropriate exclusions: no.  **Could the selection of patients have introduced bias? High**  **Applicability concerns: Low** | Index test interpreted without knowledge of reference standard: Yes  Threshold pre-specified: Yes  **Could the conduct or interpretation of the index test have introduced bias: Low**  **Applicability concerns: Low** | Reference standard likely to correctly classify the target condition: Yes  Were the reference standard results interpreted without knowledge of the results of the index test: Unclear  **Could the reference standard, its conduct, or its interpretation have introduced bias: Unclear**  **Applicability: Low** | Appropriate interval between index test and reference standard: Unclear (Surgery was done 126 (102-159) days after baseline assessment)  Did all patients receive a reference standard?No  Did patients receive the same reference standard? Yes  Were all patients included in the analysis?No  **Could the patient flow have introduced bias? High** |
| 7. Kameda 2022^7^ | Consecutive or random sample enrolled: Yes  Case-control avoided: Yes  Avoided inappropriate exclusions: Yes  **Could the selection of patients have introduced bias: Low**  **Applicability concerns: Low** | Index test interpreted without knowledge of reference standard: Yes  Threshold pre-specified: Yes  **Could the conduct or interpretation of the index test have introduced bias: Low**  **Applicability concerns: Low** | Reference standard likely to correctly classify the target condition: Yes  Were the reference standard results interpreted without knowledge of the results of the index test: Unclear  **Could the reference standard, its conduct, or its interpretation have introduced bias: Unclear**  **Applicability concerns: Low** | Appropriate interval between index test and reference standard: Unclear  Did all patients receive a reference standard? Yes  Did patients receive the same reference standard? Yes  Were all patients included in the analysis? Yes  **Could the patient flow have introduced bias? Unclear** |
| 8. vanBilsen 2022^8^ | Consecutive or random sample enrolled: Yes  Case-control avoided: Yes  Avoided inappropriate exclusions; Yes  **Could the selection of patients have introduced bias: Low**  **Applicability concerns: Low** | Index test interpreted without knowledge of reference standard: Yes  Threshold pre-specified: No;  **Could the conduct or interpretation of the index test have introduced bias: High**  **Applicability concerns: Unclear**  (CSF tap test cutoffs not mentioned for improvement) | Reference standard likely to correctly classify the target condition: Yes  Were the reference standard results interpreted without knowledge of the results of the index test: Yes  **Could the reference standard, its conduct, or its interpretation have introduced bias: Low**  **Applicability: Low** | Appropriate interval between index test and reference standard: Yes (1-3.5 months)  Did all patients receive a reference standard? Yes  Did patients receive the same reference standard? Yes  Were all patients included in the analysis? Yes  **Could the patient flow have introduced bias? Low** |
| 9. Gao 2025^9^ | Consecutive or random sample enrolled: Yes  Case-control avoided: Yes  Avoided inappropriate exclusions: No (33 patients with incomplete data and MRI excluded 12 patients were lost to follow up)  **Could the selection of patients have introduced bias: Unclear**  **Applicability concerns: Low** | Index test interpreted without knowledge of reference standard: Yes  Threshold pre-specified: Yes  **Could the conduct or interpretation of the index test have introduced bias: No**  **Applicability concerns: Low** | Reference standard likely to correctly classify the target condition: Yes  Were the reference standard results interpreted without knowledge of the results of the index test: Unclear  **Could the reference standard, its conduct, or its interpretation have introduced bias? Unclear**  **Applicability Concerns: Low** | Appropriate interval between index test and reference standard: Unclear  Did all patients receive a reference standard?No  Did patients receive the same reference standard? Yes  Were all patients included in the analysis? No  **Could the patient flow have introduced bias? High** |
| 10. Boon 1997^10^ | Consecutive or random sample enrolled: Yes  Case-control avoided: Yes  Avoided inappropriate exclusions: No (6 patients who died before the first follow up were excluded)  **Could the selection of patients have introduced bias: Unclear**  **Applicability concerns: Low** | Index test interpreted without knowledge of reference standard: Yes  Threshold pre-specified: Yes  **Could the conduct or interpretation of the index test have introduced bias: No**  **Applicability concerns: Low** | Reference standard likely to correctly classify the target condition: Yes  Were the reference standard results interpreted without knowledge of the results of the index test: Unclear  **Could the reference standard, its conduct, or its interpretation have introduced bias? Unclear**  **Applicability Concerns: Low** | Appropriate interval between index test and reference standard: Unclear  Did all patients receive a reference standard?Yes  Did patients receive the same reference standard? Yes  Were all patients included in the analysis? No  **Could the patient flow have introduced bias? High** |
| 11. Takeuchi 2000^11^ | Consecutive or random sample enrolled: Yes  Case-control avoided: Yes  Avoided inappropriate exclusions: Yes  **Could the selection of patients have introduced bias: Low**  **Applicability Concerns: Unclear** (Included only patients with atypical iNPH) | Index test interpreted without knowledge of reference standard: Yes  Threshold pre-specified: Unclear (Ro cutoff of 20 was taken; it is not clear if it was decided a priori);  **Could the conduct or interpretation of the index test have introduced bias?: Unclear**  **Applicability concerns: High** (Ro of >=20 is not standard as per literature) | Reference standard likely to correctly classify the target condition: Yes  Were the reference standard results interpreted without knowledge of the results of the index test: Unclear  **Could the reference standard, its conduct, or its interpretation have introduced bias: Unclear**  **Applicability Concerns: Low concerns** | Appropriate interval between index test and reference standard: Unclear  Did all patients receive a reference standard? Yes  Did patients receive the same reference standard? Yes  Were all patients included in the analysis? Yes  **Could the patient flow have introduced bias? Unclear** |
| 12. Sorteberg 2004^12^ | Consecutive or random sample enrolled;  Case-control avoided: Yes  Avoided inappropriate exclusions; Yes  **Could the selection of patients have introduced bias: Low**  **Applicability Concerns: Low** | Index test interpreted without knowledge of reference standard: Yes  Threshold pre-specified: Yes.  **Could the conduct or interpretation of the index test have introduced bias: Low**  **Applicability concerns: Low** | Reference standard likely to correctly classify the target condition: Unclear  Were the reference standard results interpreted without knowledge of the results of the index test: Unclear.  **Could the reference standard, its conduct, or its interpretation have introduced bias: Unclear**  **Applicability concerns: Low** | Appropriate interval between index test and reference standard: Unclear  Did all patients receive a reference standard? Yes  Did patients receive the same reference standard? Yes  Were all patients included in the analysis? Yes  **Could the patient flow have introduced bias? Unclear** |
| 13. Eide 2005^13^ | Consecutive or random sample enrolled;  Case-control avoided: Yes  Avoided inappropriate exclusions: No (Even though clinical and radiologic findings were used to select patients for shunt surgery, Rout values in 10 other subjects evaluated for possible NPH and not receiving a shunt during the same study period were lower (median value 10.5 mmHg/ml/min, ranges 6.9 – 14.5 mmHg/ml/min). There was, hence, a certain referral bias regarding preoperative Rout values and selection for shunt surgery.)  **Could the selection of patients have introduced bias? High**  **Applicability Concerns: Unclear** | Index test interpreted without knowledge of reference standard: Yes  Threshold pre-specified: Unclear (No specific mention made, but Rout 12 is the standard literature suggested threshold)  **Could the conduct or interpretation of the index test have introduced bias: Unclear**  **Applicability concerns: Low** | Reference standard likely to correctly classify the target condition: Yes  Were the reference standard results interpreted without knowledge of the results of the index test: Unclear  **Could the reference standard, its conduct, or its interpretation have introduced bias: Unclear**  **Applicability concerns: Low** | Appropriate interval between index test and reference standard: Yes (A ventriculo-peritoneal (VP) shunt was implanted 2 - 3 weeks)  Did all patients receive a reference standard? Yes  Did patients receive the same reference standard? Yes  Were all patients included in the analysis? Yes  **Could the patient flow have introduced bias? Low** |
| 14. Hasselbalch 2023^14^ | Consecutive or random sample enrolled: yes  Case-control avoided: Yes  Avoided inappropriate exclusions: Unclear, "361 consecutive patients diagnosed  with possible or probable iNPH. 127 patients met the inclusion criteria for; Only patients who had outcomes of at least 2 months were included. Out of 361, only 127 patients were included in the analysis) (Unclear if all clinico-radiologically diagnosed patients were selected to be enrolled in the study)  **Could the selection of patients have introduced bias: Unclear**  **Applicability concerns: Unclear** | Index test interpreted without knowledge of reference standard: Yes  Threshold pre-specified: No  **Could the conduct or interpretation of the index test have introduced bias: High**  **Applicability concerns: Low** | Reference standard likely to correctly classify the target condition: No (Due to the retrospective design of the study, post-operative objective measures for gait and cognition (10-m gait test and MMSE) were only available in 70% and 80% of the patients, respectively.  Were the reference standard results interpreted without knowledge of the results of the index test: Unclear.  **Could the reference standard, its conduct, or its interpretation have introduced bias? High**  **Applicability Concerns: High** | Appropriate interval between index test and reference standard: Unclear  Did all patients receive a reference standard?No  Did patients receive the same reference standard? Yes  Were all patients included in the analysis?No  **Could the patient flow have introduced bias? High** |
