## Supplementary table 4 for "Accuracy of CSF Tap Test and Lumbar Infusion Test in Predicting Shunt Response in Idiopathic Normal Pressure Hydrocephalus: A Systematic Review and Meta-Analysis"

**Question**: Should CSF Tap test be used to screen for shunt responsiveness in idiopathic Normal Pressure hydrocephalus?

| Sensitivity | 0.68 (95% CI: 0.52 to 0.80) |
| --- | --- |
| Specificity | 0.53 (95% CI: 0.41 to 0.66) |

| Prevalences | 50% | 60% | 80% |
| --- | --- | --- | --- |

| Outcome | № of studies (№ of patients) | Study design | Factors that may decrease certainty of evidence | | | | | Effect per 1,000 patients tested | | | Test accuracy CoE |
| --- | --- | --- | --- | --- | --- | --- | --- | --- | --- | --- | --- |
|  |  |  | Risk of bias | Indirectness | Inconsistency | Imprecision | Publication bias | pre-test probability of 50% | pre-test probability of 60% | pre-test probability of 80% |  |
| **True positives** (patients with shunt responsiveness) | 9 studies 697 patients | cross-sectional (cohort type accuracy study) | serious^a^ | not serious | very serious^b^ | serious^c^ | none | 338 (261 to 399) | 405 (313 to 479) | 540 (418 to 638) | ⨁◯◯◯ Very low^a,b,c^ |
| **False negatives** (patients incorrectly classified as not having shunt responsiveness) |  |  |  |  |  |  |  | 162 (101 to 239) | 195 (121 to 287) | 260 (162 to 382) |  |
| **True negatives** (patients without shunt responsiveness) | 9 studies 697 patients | cross-sectional (cohort type accuracy study) | serious^a^ | not serious | serious^d^ | not serious^e^ | none | 267 (204 to 328) | 213 (163 to 262) | 107 (81 to 131) | ⨁⨁◯◯ Low^a,d,e^ |
| **False positives** (patients incorrectly classified as having shunt responsiveness) |  |  |  |  |  |  |  | 233 (172 to 296) | 187 (138 to 237) | 93 (69 to 119) |  |

#### Explanations

a. High risk of bias for patient selection and flow and timing domains

b. Point estimates are all over the place, High statistical Heterogeneity

c. The confidence intervals are crossing a clinically meaningful threshold of 70% sensitivity, but the upper limit of the confidence interval is 80%, showing possible usefulness

d. Considering a clinically meaningful specificity threshold of 70%, most studies fall below this threshold with moderate statistical heterogeneity

e. Considering a specificity of 70% to be clinically meaningful, both sides of the confidence interval are below this threshold, thus providing a precise result that the test poorly specific
