## Appendix 1 for "Accuracy of CSF Tap Test and Lumbar Infusion Test in Predicting Shunt Response in Idiopathic Normal Pressure Hydrocephalus: A Systematic Review and Meta-Analysis"

### Pubmed/Web of Science/Scopus

#1 (((normal pressure hydrocephalus) OR (NPH)) OR (iNPH)) OR (idiopathic normal pressure

hydrocephalus)

#2 (((CSF tap test) OR (large volume CSF tap) OR (magnetic resonance imaging)) OR (MRI)) OR

(lumbar infusion test)

#3 Ventriculoperitoneal shunt OR ventriculo peritoneal shunt OR shunt surgery OR CSF diversion procedure OR lumbar drain OR lumboperitoneal shunt OR thecoperitoneal shunt OR theco peritoneal shunt

((#1) AND (#2)) AND (#3)

### Cochrane central

#1 Normal Pressure Hydrocephalus

#2 Idiopathic Normal Pressure Hydrocephalus

#3 NPH

#4 iNPH

#5 #1 OR #2 OR #3 OR #4

#6 CSF tap test

#7 Lumbar infusion test

#8 large volume CSF tap

#9 MRI

#10 Magnetic Resonance Imaging

#11 #6 OR #7 OR #8 OR #9 OR #10

#12 Ventriculoperitoneal shunt

#13 Ventriculo peritoneal shunt

#14 Shunt surgery

#15 CSF diversion procedure

#16 lumbar drain

#17 lumboperitoneal shunt

#18 thecoperitoneal shunt

#19 theco peritoneal shunt

#20 #12 OR #13 OR #14 OR #15 OR #16 OR #17 OR #18 OR #19

#21 #5 AND #11 AND #20

### EMBASE

#1 Normal Pressure Hydrocephalus

#2 Idiopathic Normal Pressure Hydrocephalus

#3 NPH

#4 iNPH

#5 #1 OR #2 OR #3 OR #4

#6 CSF tap test

#7 Lumbar infusion test

#8 large volume CSF tap

#9 MRI

#10 Magnetic Resonance Imaging

#11 #6 OR #7 OR #8 OR #9 OR #10

#12 Ventriculoperitoneal shunt

#13 Ventriculo peritoneal shunt

#14 Shunt surgery

#15 CSF diversion procedure

#16 lumbar drain

#17 lumboperitoneal shunt

#18 thecoperitoneal shunt

#19 theco peritoneal shunt

#20 #12 OR #13 OR #14 OR #15 OR #16 OR #17 OR #18 OR #19

#21 #5 AND #11 AND #20
