## Supplementary figures and images for "Accuracy of CSF Tap Test and Lumbar Infusion Test in Predicting Shunt Response in Idiopathic Normal Pressure Hydrocephalus: A Systematic Review and Meta-Analysis"

### Supplementary figure 1

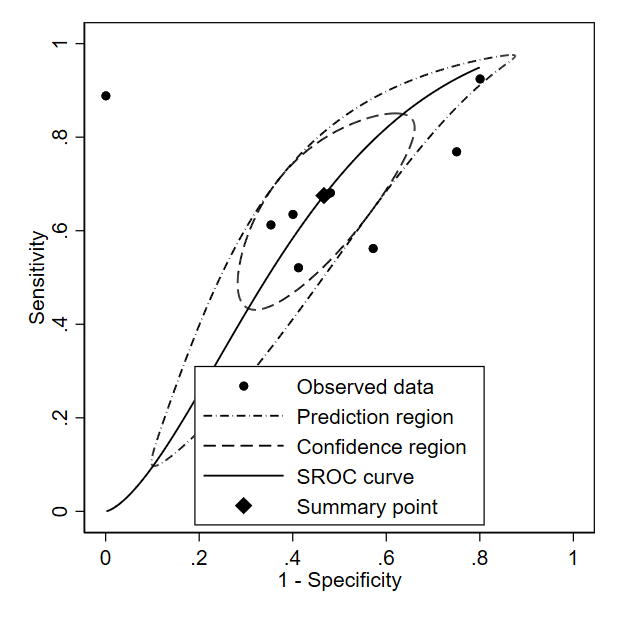

### Supplementary figure 2

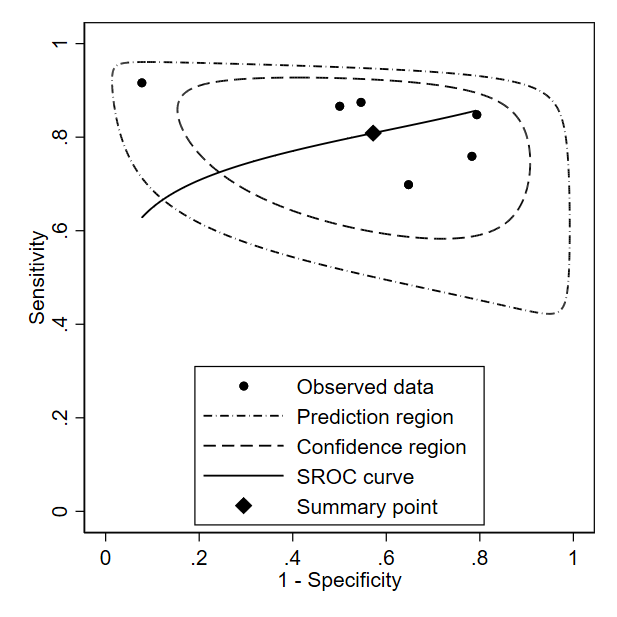
